# Clinical correlations of SARS-CoV-2 antibody responses in patients with COVID-19 infection

**DOI:** 10.1101/2020.10.22.20213207

**Authors:** Mia DeSimone, Daimon P Simmons, Nicole Tolan, Stacy Melanson, Athena Petrides, Milenko Tanasijevic, Peter Schur

## Abstract

Coronavirus disease 19 (COVID-19) is caused by the severe acute respiratory syndrome coronavirus 2 (SARS-CoV-2). Understanding the clinical correlations of antibodies produced by infected individuals will be critical for incorporating antibody results into clinical management. This study was an observational cohort study to evaluate antibody responses in individuals with PCR-confirmed COVID-19, including 48 hospitalized patients diagnosed with COVID-19 by real-time polymerase chain reaction (RT-PCR) at a large tertiary care medical center. Serum samples were obtained from patients at various time points during the disease course and tested for IgM and IgG antibodies against SARS-CoV-2. Medical records were reviewed, and antibody levels were compared with clinical and laboratory findings. Patients did not have high levels of antibodies within one week of symptoms, but most had detectable IgM and IgG antibodies between 8 and 29 days after onset of symptoms. Some individuals did not develop measurable levels of IgM or IgG antibodies. IgM antibodies were associated with elevated ALT, but there were no other significant associations. We did not observe significant associations of SARS-CoV-2 antibodies with clinical outcomes, including intubation and death. SARS-CoV-2 IgM and IgG antibodies were unlikely to be detected in the first week of infection or in severely immunocompromised individuals. Although we did not observe associations with clinical outcomes, IgM antibodies were associated with higher ALT levels. Antibody production reflects the virus-specific immune response, which is important for immunity but also drives pathology, and antibody levels may be important for guiding treatment of individuals with COVID-19.

## Introduction

Coronavirus disease 19 (COVID-19) is caused by the severe acute respiratory syndrome coronavirus 2 (SARS-Cov-2) and is currently leading to a major global pandemic after originating in Wuhan, China in late 2019.^1–11^ Individuals infected with this virus have a range of symptoms including cough, difficulty breathing, fever, chills, muscle pain, headache, sore throat, loss of smell or taste, and gastrointestinal symptoms.^1,5–15^ Many individuals with COVID-19 are asymptomatic or report mild symptoms but others, especially those of advanced age with underlying comorbidities, can become critically ill with respiratory failure, septic shock, and/or multiorgan dysfunction or failure.^5–8,10–12,14–16^ Although immune responses are critical for protection against the virus, significant mortality results from immune activation and cytokine storm, and immune suppression may paradoxically play a critical role in the treatment of patients with complications from excess immune activation.^17,18^

SARS-CoV-2 is an enveloped, single-stranded RNA virus of the family *Coronaviridae* that contains four structural glycoproteins: envelope (E), membrane (M), nucleocapsid (N), and spike (S).^3,4^ Detection of SARS-CoV-2 viral RNA by reverse transcriptase-polymerase chain reaction (RT-PCR) in samples collected from nasopharyngeal swabs or saliva is the gold standard diagnostic test to confirm early infection.^7,8,15,19^ The sensitivity of these diagnostic tests is limited by the adequacy of the sample collection, and viral RNA may only be detectable during the acute phase of infection.^15,19,20^ Detection of antibodies may further elucidate the immune status of infected individuals ^20–26^ and improve the sensitivity of identifying patients with COVID-19.^21^ In severe acute respiratory syndrome (SARS), virus-specific antibodies were detectable in 80-100% of patients two to three weeks after symptom onset.^27–29^ Initial reports have suggested that SARS-CoV-2-specific IgM is detectable in the acute phase and within one to two weeks of infection, whereas virus-specific IgG takes two to four weeks to become detectable during the recovery phase.^20–26,30^ Many assays have become available to detect antibodies against SARS-CoV-2,^31^ with differing sensitivity and specificity.^32,33^

Patients may develop neutralizing antibodies against SARS-CoV-2 that will be important for immunity.^34,35^ However, higher viral-specific antibody levels have also been associated with a worse clinical prognosis.^21^ Consistent with this discrepancy, in animal models of infection with other related coronaviruses, antibodies contribute to inflammation and pathology,^36–39^ and immune activation is an important consideration for SARS-CoV-2.^40^ The balance of protection and inflammation driven by specific antibodies to SARS-CoV-2 is not yet well understood. To address this gap in knowledge, we performed a detailed analysis of serological and clinical results for a cross-section of patients in a large tertiary academic medical center during the COVID-19 pandemic.

## Methods

### Patient Selection

Samples were collected as discarded specimens from routine clinical care of forty-eight patients diagnosed with COVID-19 and hospitalized at Brigham and Women’s Hospital in Boston, MA from April 3 to April 17, 2020. Patients with COVID-19 were identified based on a previous positive lab result for SARS-CoV-2 nucleic acid by RT-PCR using nasopharyngeal swab samples. There were no other specific inclusion or exclusion criteria. Many patients were discharged shortly after initial diagnosis, and serial serum samples were obtained from 17 patients over this 14-day period. Medical records were reviewed for past medical history, date of illness onset, presenting symptoms, hospital length of stay, and outcome. Laboratory values were extracted if available within 48 hours of the corresponding serum sample used for this analysis.

### Reagents/Antibody Measurement

IgM and IgG antibodies against SARS-CoV-2 were detected in serum samples using enzyme-linked immunosorbent assay (ELISA) kits (Epitope Diagnostics, San Diego, CA) and performed according to the manufacturer’s instructions. This assay uses nucleocapsid antigens to detect IgM and IgG antibodies that bind SARS-CoV-2. The manufacturer’s reported sensitivity is 98.4% for IgG and 73.8% for IgM. The manufacturer’s reported specificity is 99.8% for IgG and 100% for IgM. This assay had similar performance in our clinical validation and in additional studies performed in our lab.^41^ Arbitrary units for SARS-CoV-2 IgM and IgG were calculated based on the optical density of the sample normalized to the positive cut-off defined by the optical density of the negative control. The positive cut-off was set to a value of 1, and samples with units between 0.82 and 1 were reported as borderline. For our analysis, borderline results were treated as negative.

### Data Analysis

The last sample for patients with serial samples was selected for analysis of unique patients. Nonparametric statistical analysis (Mann-Whitney) was used for laboratory findings, with values at the minimum or maximum measurements counted as ties. Patients with unavailable data for a laboratory test were excluded from the analysis for that test, as indicated in the table. Graphical and statistical analysis was performed in Microsoft Excel 16.36 (Microsoft) and Prism 6.0.h (GraphPad). P.S., N.T., S.M., A.P., M.T., D.P.S., and M.D participated in study design, interpretation, and writing. Data analysis was performed by D.P.S. and M.D.

### Ethical Considerations

This study was reviewed and approved by the Brigham and Women’s Hospital Institutional Review Board (Boston, MA, USA).

## Results

### Patient Characteristics

In order to study antibody responses to SARS-CoV-2, we identified 48 hospitalized patients at our institution who had at least one previous positive SARS-CoV-2 RT-PCR test and serum for additional testing. Serum samples were collected over the course of two weeks (April 3 to April 17, 2020) to provide a cross-section of the SARS-CoV-2 serology responses within the hospital. The cohort of patients included in the study were individuals with diverse demographics, presenting symptoms, and comorbidities (Table 1). This cohort included a broad spectrum of individuals within the hospital: 52% women and 48% men from a variety of races and ethnicities, including 48% Black or African American, 29% Hispanic or Latino, and 27% White. Comorbidities were present in the vast majority (92%) of patients, with a high prevalence of hypertension (48%), prior or current malignancy (29%), and diabetes (27%). Patients reported a wide range of symptoms that led them to first present to our institution for medical attention. All but two individuals in this cohort (96%) endorsed more than one presenting symptom. Cough (81%), fever (73%), and shortness of breath (58%) were commonly reported by these individuals.

**Table 1.** Characteristics of the Patients with SARS-CoV-2 Infection at Presentation

|  |  |
| --- | --- |
| Characteristic | N = 48 |
| Median age (IQR) -- yr. | 61 (49-69) |
| Female sex -- no. (%) | 25 (52) |
| Race/ethnicity -- no. (%) |  |
| Black or African American | 16 (48) |
| Hispanic or Latino | 14 (29) |
| White, non-Hispanic | 13 (27) |
| Asian | 1 (2) |
| Other or not specified | 4 (8) |
| Comorbidities -- no. (%) |  |
| Cardiovascular disease | 7 (15) |
| Cerebrovascular disease | 2 (4) |
| Chronic lung disease | 8 (17) |
| Diabetes | 13 (27) |
| Hypertension | 23 (48) |
| Malignancy* | 14 (29) |
| Other chronic illness | 28 (58) |
| None | 4 (8) |
| Clinical criteria -- no. (%)† |  |
| Chills | 10 (21) |
| Cough | 39 (81) |
| Fever | 35 (73) |
| Headache | 10 (21) |
| Muscle pain | 13 (27) |
| New loss of taste or smell | 5 (10) |
| Shortness of breath | 28 (58) |
| Sore throat | 8 (17) |
| Repeated shaking with chills | 2 (4) |
| Other | 29 (60) |
| Gastrointestinal symptoms‡ | 20 (42) |
| Cardiac symptoms§ | 5 (10) |
| *Prior or current malignancy |  |
| †Clinical criteria based on the CDC case definition of COVID-19 |  |
| ‡Nausea, vomiting, diarrhea, abdominal pain and/or anorexia |  |
| §Chest pain or chest tightness |  |

### High levels of SARS-CoV-2 IgM and IgG antibodies detected in a subset of patients

We evaluated IgM and IgG antibody responses to SARS-CoV-2 in patients over the course of infection based on reported onset of symptoms extracted from the medical record (median 14 days, interquartile range 11-18 days). We did not detect high levels of IgM or IgG antibodies in any samples collected from patients within the first seven days of infection (Fig 1A-B). The highest IgM levels (greater than 10 units) were detected from 10 to 16 days after onset of symptoms (Fig 1A). In contrast, a wide range of IgG levels were measured throughout the course of infection, including positive results from 8 to 29 days after onset of symptoms (Fig 1B). In addition, nine individuals did not develop high IgM or IgG antibodies against SARS-CoV-2 within this time frame. We analyzed serial samples to determine whether this finding might be due to rising or falling levels of IgM and IgG over time. IgM levels rose in six patients at early time points and were stable at later time points (Fig 1C). In contrast, samples from eleven patients collected over various time points showed increasing levels of IgG antibody (Fig 1D).

**Figure 1.**
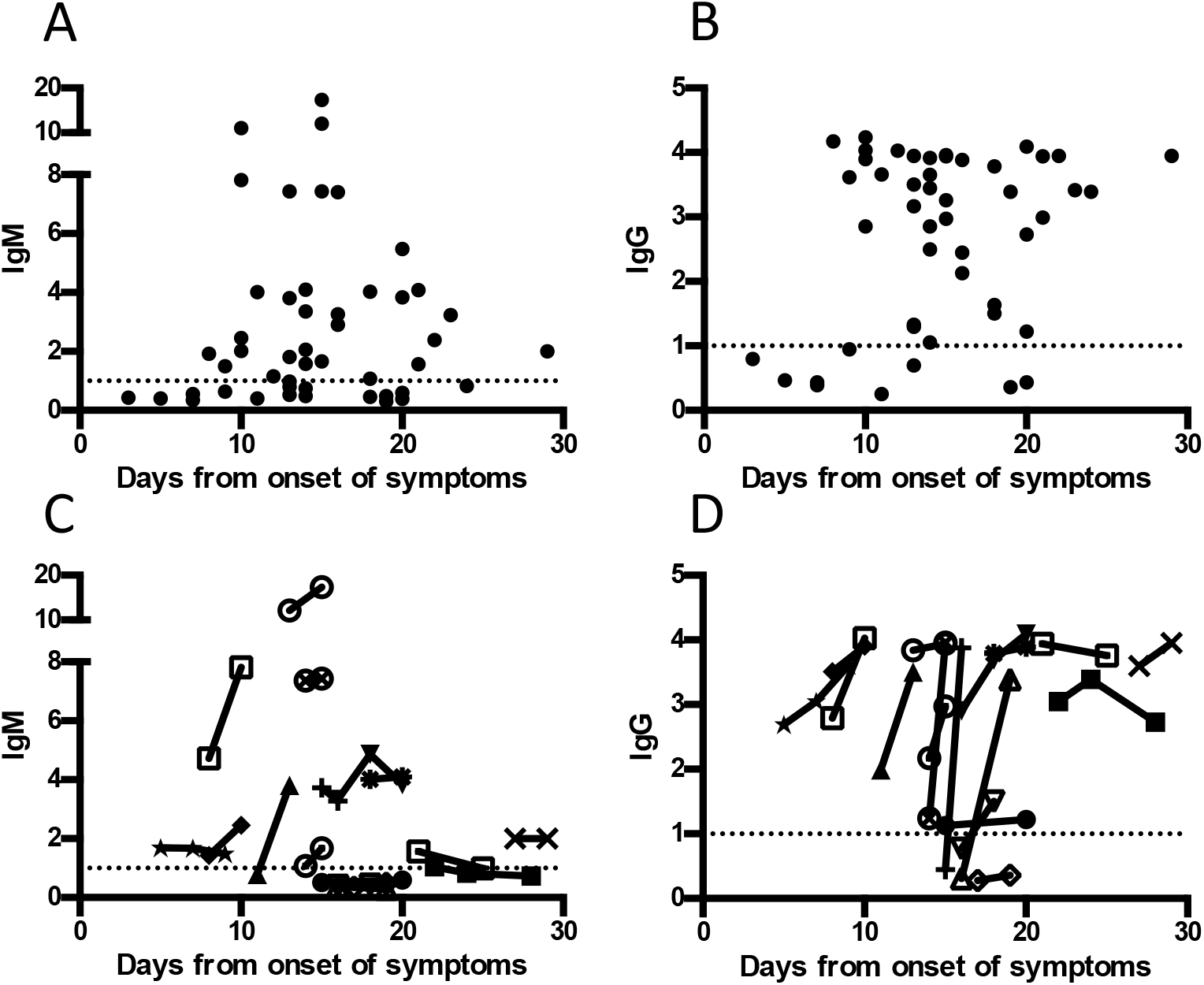
IgM and IgG antibody responses over the course of infection. A, IgM for individual patients relative to reported onset of symptoms. The sample from the latest time point is shown for individuals with more than one sample. B, IgG for individual patients relative to reported onset of symptoms. The sample from the latest time point is shown for individuals with more than one sample. C, IgM levels for patients with serial samples relative to reported onset of symptoms. Measurements from the same patient are drawn connected by a line. D, IgG levels for patients with serial samples relative to reported onset of symptoms. Measurements from the same patient are drawn connected by a line. The reference range is negative <0.820, borderline 0.820-0.999, positive >0.999.

To control for the subjective nature of symptom reporting, we analyzed antibody results relative to the date of the first positive SARS-CoV-2 RT-PCR result, as a secondary measure of the time of acute infection. The highest IgM levels (greater than 10 units) were detected within 10 days (median 7 days, interquartile range 6-8 days) from the first positive SARS-CoV-2 RT-PCR, with very low levels detected at later time points (Fig 2A). In contrast, IgG levels varied over time relative to the first positive SARS-CoV-2 RT-PCR result (median 5 days, interquartile range 2-8 days) (Fig 2B). In the analysis of serial samples, IgM levels rose within the first week after a positive SARS-CoV-2 RT-PCR result and remained stable at later time points (Fig 2C). Similarly, IgG levels for all but one of the patients rose within the first week after the first positive SARS-CoV-2 RT-PCR result, and three patients had stable levels of IgG more than 15 days after the positive RT-PCR (Fig 2D). Notably, nine individuals had persistently low levels of IgM and IgG antibodies, even long after initial infection defined both by symptoms and by detection of viral RNA.

**Figure 2.**
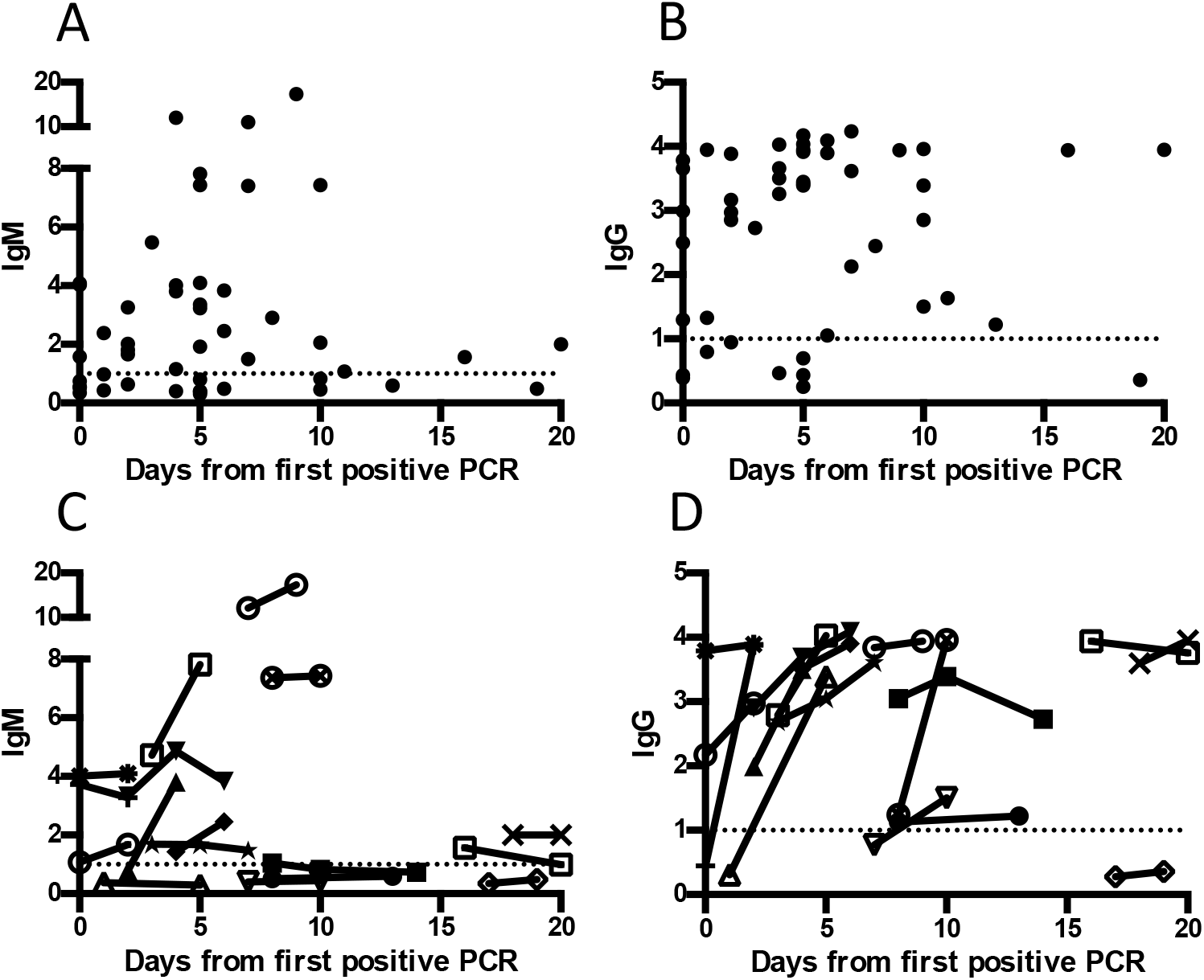
IgM and IgG antibody responses relative to positive RT-PCR. A, IgM for individual patients relative to the first positive COVID RT-PCR. The sample from the latest time point is shown for individuals with more than one sample. B, IgG for individual patients relative to the first positive COVID RT-PCR. The sample from the latest time point is shown for individuals with more than one sample. C, IgM levels for patients with serial samples relative to the first positive COVID RT-PCR. Measurements from the same patient are drawn connected by a line. D, IgG levels for patients with serial samples relative to the first positive COVID RT-PCR. Measurements from the same patient are drawn connected by a line. The reference range is negative <0.820, borderline 0.820-0.999, positive >0.999.

### Characteristics of patients with very low levels of SARS-CoV-2 IgM and IgG antibodies

We performed detailed chart reviews to identify clinical findings of patients who did not develop high levels of IgM or IgG antibodies against SARS-CoV-2 (Table 2). Samples from two individuals were collected less than one week after reported onset of symptoms, and it is possible that upon subsequent testing, antibodies would have been identified. Antibodies were also not detected in samples from three additional patients collected within two weeks of reported onset of symptoms. Although the low antibody levels could be attributed to early sample collection for six patients, two individuals did not produce high levels of SARS-CoV-2 antibodies even later in the course of infection. One individual with B cell acute lymphoblastic leukemia developed cough and headache while hospitalized for induction chemotherapy. This patient tested positive for SARS-CoV-2 by RT-PCR but did not develop SARS-CoV-2 antibodies as late as 19 days after onset of symptoms. Another patient with acute myeloid leukemia was admitted to the hospital with neutropenic fever and found to be positive for SARS-CoV-2 by RT-PCR but had undetectable levels of SARS-CoV-2 antibodies as far out as 20 days from onset of symptoms. In contrast, five patients on immunosuppressive therapies did develop IgG antibodies, of which three also had IgM antibodies. including a patient post-transplant for myelofibrosis, on ruxolitinib and prednisone. Two patients produced SARS-CoV-2 antibodies despite a recent history of chemotherapy (alectinib, high-dose cytarabine, and etoposide for one patient, carboplatin/gemcitabine for another). Another patient with detectable SARS-CoV-2 antibodies was started on prednisone while in the hospital. These data indicate that, even with immunosuppressive medications, most hospitalized patients will develop antibodies against SARS-CoV-2 unless they are profoundly immunosuppressed.

**Table 2.**
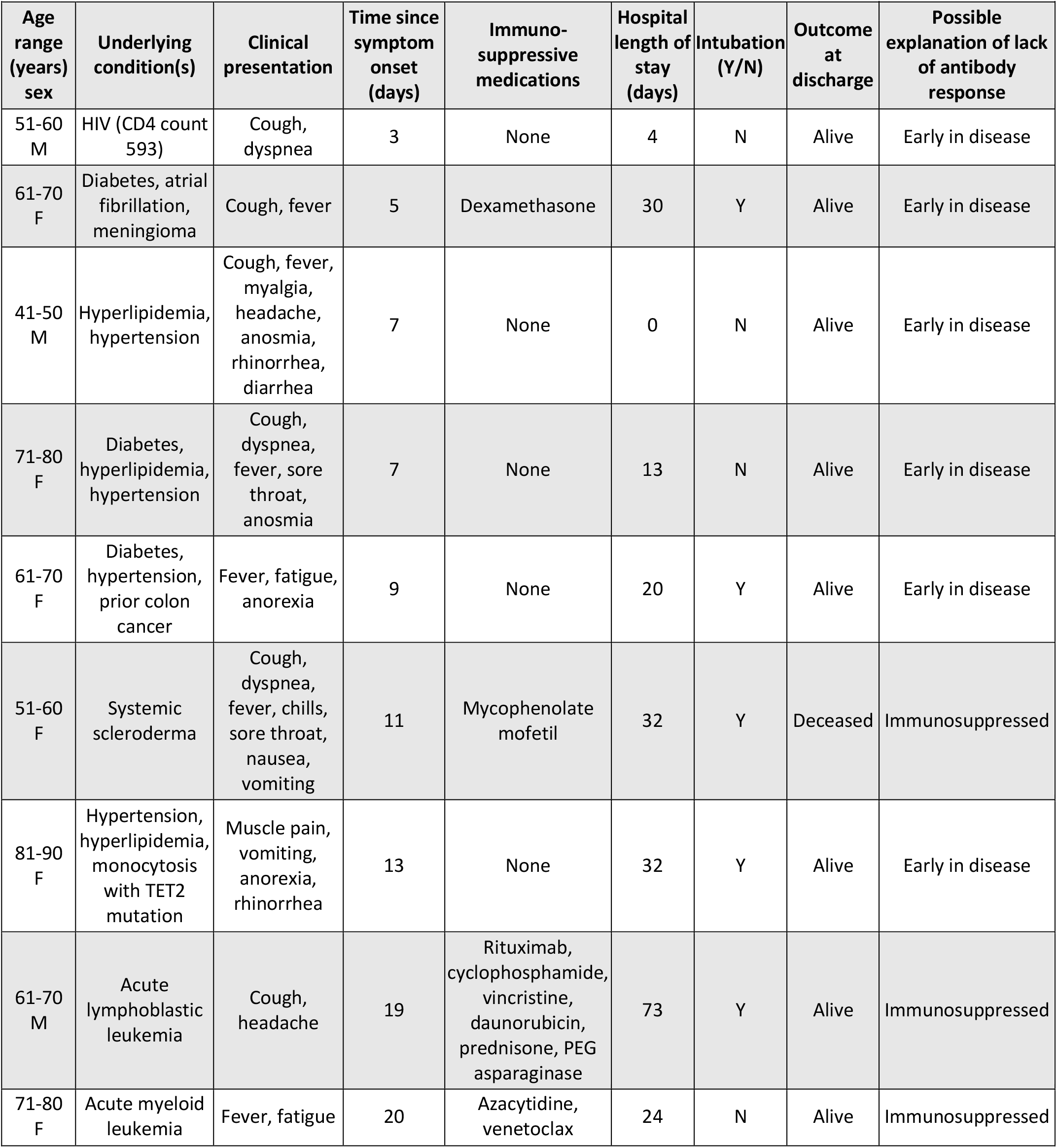
Clinical characteristics of patients infected with SARS-CoV-2 with low or undetectable SARS-CoV-2 IgM and IgG antibodies

### Association of SARS-CoV-2 IgM and IgG antibodies with other laboratory findings

In order to identify other correlates of antibody responses, we evaluated the association of SARS-CoV-2 antibodies with other laboratory findings. We analyzed other laboratory results for patients with (n=30) or without (n=18) SARS-CoV-2 IgM antibodies (Table 3). The individuals without IgM antibodies were 61% female, 33% Black or African American, 33% Hispanic or Latino, and 28% White, compared to 47% female, 33% Black or African American, 27% Hispanic or Latino, and 27% White in the group positive for IgM antibodies. The median age was 65 (interquartile range 54-71 years) among IgM-negative individuals and 52 (interquartile range 47-67 years) among IgM-positive individuals. Individuals with IgM had higher ALT levels (Table 3). We did not observe an association between lymphocyte count and IgM levels. We also did not find significant associations of SARS-CoV-2 IgM antibodies with AST, globulin, troponin, ferritin, D-dimer, procalcitonin, or platelet count levels. The hospital stay was shorter for IgM-positive individuals (median 8 days, interquartile range 5.5-18.5 days) than IgM-negative individuals (median 13 days, interquartile range 4-24 days). The rate of intubation was 56% (n=10) in IgM-negative individuals versus 50% (n=15) in those with detectable IgM antibodies. The rate of death was 17% (n=3) in the IgM-negative group versus 13% (n=4) in the IgM-positive group for the duration of this study.

**Table 3.** Correlation of SARS-CoV-2 IgM responses to other laboratory findings

|  |  |  |  |  |
| --- | --- | --- | --- | --- |
| **Result\*** | **IgM negative median (IQR)** | **IgM positive median (IQR)** | **Mann-Whitney p value** | **Bonferroni adjusted p value** |
| ALT (U/mL) | 22 (17, 35) | 42 (21, 61) | 0.003 | 0.029 |
| AST (U/mL) | 25 (16, 52) | 46 (33, 85) | 0.083 | 0.751 |
| D-dimer (ng/mL) | 854 (540, 1855) | 1990 (893, 3900) | 0.065 | 0.587 |
| Ferritin (ug/dL) | 606 (450, 1103) | 1309 (742, 1712) | 0.062 | 0.554 |
| Globulin (g/dL) | 3 (3, 3) | 3.4 (3.1, 3.8) | 0.017 | 0.149 |
| Lymphocyte count (K/uL) | 1.1 (0.7, 1.6) | 1 (0.8, 1.7) | 0.935 | 1.000 |
| Platelet count (K/uL) | 223 (161, 247) | 264 (190, 354) | 0.278 | 1.000 |
| Procalcitonin (ng/mL) | 0.17 (0.08, 0.42) | 0.25 (0.13, 0.59) | 0.368 | 1.000 |
| Troponin T (ng/L) | 12 (8, 23) | 9 (6, 22) | 0.558 | 1.000 |
| *Laboratory values were extracted from IgM negative (n=18) and IgM positive (n=30) individuals. Each laboratory value was available for at least 72% of IgM negative individuals and at least 67% of IgM positive individuals. |  |  |  |  |

We performed a similar analysis of laboratory results for patients with (n=39) or without (n=9) SARS-CoV-2 IgG antibodies (Table 4). Individuals without IgG antibodies were 67% female, 33% Black or African American, 33% Hispanic or Latino, and 22% White. Those with IgG antibodies were 49% female, 33% Black or African American, 28% Hispanic or Latino, and 28% White. The median age was 64 (interquartile range 55-71 years) for IgG-negative individuals and 54 (interquartile range 47-69 years) for IgG-positive individuals. There were no significant associations of laboratory results with IgG levels, although there was a trend towards higher ALT, globulin, and D-dimer levels that was not statistically significant after correcting for multiple comparisons (Table 4). The hospital stay was shorter for IgG-positive individuals (median 8 days, interquartile range 5.25-18.75 days) compared with IgG-negative individuals (median 20 days, interquartile range8.5-28 days). The rate of intubation was 56% (n=5) among IgG-negative individuals and 51% (n=20) among IgG-positive individuals. There was one fatality (11%) among the patients without IgG antibodies and six fatalities (15%) among the patients with detectable IgG antibodies for the duration of this study.

**Table 4.** Correlation of SARS-CoV-2 IgG responses to other laboratory findings

|  |  |  |  |  |
| --- | --- | --- | --- | --- |
| **Result\*** | **IgG negative median (IQR)** | **IgG positive median (IQR)** | **Mann-Whitney p value** | **Bonferroni adjusted p value** |
| ALT (U/mL) | 22 (19, 34) | 34 (20, 61) | 0.082 | 0.742 |
| AST (U/mL) | 25 (18, 40) | 44 (27, 76) | 0.580 | 1.000 |
| D-dimer (ng/mL) | 775 (501, 1815) | 1840 (865, 3637) | 0.049 | 0.440 |
| Ferritin (ug/dL) | 616 (351, 1153) | 1228 (588, 1689) | 0.345 | 1.000 |
| Globulin (g/dL) | 2.8 (2.7, 3.3) | 3.4 (2.8, 3.6) | 0.100 | 0.900 |
| Lymphocyte count (K/uL) | 1.1 (0.7, 1.3) | 1.1 (0.8, 1.7) | 0.397 | 1.000 |
| Platelet count (K/uL) | 210 (145, 242) | 239 (190, 348) | 0.174 | 1.000 |
| Procalcitonin (ng/mL) | 0.26 (0.09, 0.61) | 0.25 (0.1, 0.5) | 0.962 | 1.000 |
| Troponin T (ng/L) | 12 (10, 19) | 9 (6, 24) | 0.576 | 1.000 |
| *Laboratory values were extracted from IgG negative (n=9) and IgG positive (n=39) individuals. Each laboratory value was available for at least 78% of IgG negative individuals and at least 69% of IgM positive individuals. |  |  |  |  |

## Discussion

In this study of hospitalized patients infected with SARS-CoV-2, we found that most patients had detectable SARS-CoV-2 IgM and IgG antibodies after a week following symptom onset, ranging from 8 to 29 days. Importantly, we only detected very low levels of IgM or IgG within the first week of infection. In this cohort of patients with confirmed COVID-19, nine out of forty-eight individuals did not produce high levels of IgM during the course of infection. This pattern might represent a less robust early immune response by individuals with modest symptoms. Thirty-nine out of forty-eight individuals developed IgG antibodies against SARS-CoV-2. The fact that we observed IgG antibodies more frequently than IgM might suggest a key role for helper T cells in generating antibody responses at later time points in infection. However, we did observe a modest association of IgM antibodies with elevated ALT enzymes. In addition to helping diagnose acute infection, high levels of IgM may represent a phase of the acute immune activation that can lead to disease pathology.

Although low antibody levels could be attributed to sampling early in the course of infection for some individuals, others did not have elevated antibodies even at later time points. Three severely immunocompromised individuals did not produce antibody responses, and this is an important consideration in immunocompromised patients for both diagnosis and monitoring potential protective immunity to future infection. Although these three severely immunocompromised patients did not produce high levels of antibodies, five individuals receiving immunosuppressive medications did have detectable IgG and/or IgM antibodies. This discrepancy might be explained by differences in the timing of immunosuppression or could represent alternative functions of different classes of immunosuppressive medications. Although immunosuppressed individuals are at risk for not developing antibodies against SARS-CoV-2, many will develop antibodies unless profoundly immunosuppressed. This assay measures nucleocapsid antibodies, whereas antibodies against the receptor binding domain of the spike protein have been correlated with viral neutralization.^34,35^ However, it is currently unclear whether SARS-CoV-2 antibodies confer immunity,^35^ and if so, what titers against different viral antigens would be necessary. Additional studies will be essential to address this question.

This study provides new information about antibody responses to SARS-CoV-2 in a U.S. population, with important clinical information about individuals who do not develop high levels of SARS-CoV-2 IgM or IgG antibodies. However, this study centers on a small population focused at a single tertiary care, large academic medical center and is based on the experience with a single serology assay. Very few individuals remained hospitalized at later time points in infection, so it was not feasible to obtain a large number of samples from late time points. Lack of follow-up after discharge limits the ability to identify possible correlations with long-term morbidity. Furthermore, other serology assays may feature improved sensitivity, particularly in the early detection of SARS-CoV-2 antibodies. Additional studies with larger numbers of patients and multiple centers will be necessary to more definitively determine the clinical status of patients with low and high levels of SARS-CoV-2 IgM and IgG antibodies and to better define how these antibody levels relate to clinical outcomes.

## Data Availability

No large datasets have been generated in this manuscript.

## Acknowledgments

We thank Lisa Bernhard and Annmarie Anderson for testing samples, and Mike Conrad for assistance identifying samples.

